# Predicting 5-Year Breast Cancer Risk from Longitudinal Digital Breast Tomosynthesis: A Single-center Retrospective Study

**DOI:** 10.64898/2026.03.22.26349001

**Authors:** Yanqi Xu, Laura Heacock, Jungkyu Park, Felicia L Pasadyn, Qi Lei, Alana Lewin, Krzysztof Jerzy Geras, Linda Moy, Freya Schnabel, Yiqiu Shen

## Abstract

**Key Results:** In an independent test set, a longitudinal DBT-based deep learning model achieved higher 5-year risk discrimination than the FFDM-based Mirai model (AUC, 0.720 vs 0.687; p < 0.001) and, in a matched case-control cohort, higher discrimination than the Tyrer–Cuzick model (AUC, 0.676 vs 0.567; p < 0.001).

**Importance:** Longitudinal DBT-based AI improves individualized breast cancer risk assessment beyond FFDM-based AI and clinical risk models.

**Background:** Imaging-based breast cancer risk prediction models primarily use full-field digital mammography (FFDM). As digital breast tomosynthesis (DBT) has become a predominant screening modality in the United States, its potential for long-term breast cancer risk prediction remains under-explored.

**Objective:** To develop and evaluate a deep learning model that uses longitudinal DBT exams to predict long-term breast cancer risk.

**Methods:** This retrospective study included 313,531 DBT exams from 161,165 women (mean age, 58.5 ± 11.7 years) between January 2016 and August 2020 at Institute A. A risk prediction (DRP) model was developed to estimate 2–5 year breast cancer risk using longitudinal DBT exams, patient age and breast density. Model performance was compared with a single-time point DBT model, the Mirai model using same-day FFDM, and the Tyrer-Cuzick model using the area under the receiver operating characteristic curve (AUC), time-dependent concordance index, and integrated Brier score.

**Results:** In an independent test set (n = 34,580), the longitudinal DRP model achieved a 5-year AUC of 0.720 (95% CI, 0.703–0.738), improving on the single time point DRP model (AUC, 0.706; 95% CI, 0.687–0.724; p < 0.001) and the Mirai model (AUC, 0.687; 95% CI, 0.668–0.705; p < 0.001). In a matched case-control cohort (n=432), the DRP model achieved a 5-year AUC of 0.676 (95% CI, 0.626–0.727), compared with 0.567 (95% CI, 0.514–0.621; p < 0.001) for the Tyrer-Cuzick model. The model reclassified 37.6% (705/1,877) of women with extremely dense breasts as average risk, with a 5-year cancer incidence of 0.7% (5/705), and identified 15.5% (404/2,605) of women with fatty breasts as high risk, with a 5-year cancer incidence of 2.5% (10/404).

**Conclusion:** A deep learning model using longitudinal DBT examinations improved long-term breast cancer risk prediction compared with FFDM-based and clinical risk models.

**Clinical Impacts:** Longitudinal DBT-based risk prediction may enable dynamic risk assessment using screening images, supporting personalized screening strategies and more targeted use of supplemental imaging.

## Introduction

Breast cancer is the second most common cancer among women in the U.S. and the leading cause of cancer-related deaths among women worldwide.^1,2^ While screening aims to detect existing malignancy, risk prediction estimates the likelihood that a cancer-free individual will develop cancer in the future. Accurate risk prediction is central to preventive care, guiding decisions regarding screening intervals, supplemental imaging, and risk-reduction strategies. As screening programs are shifting toward risk-based approaches,^3^ reliable individualized risk estimates are needed to balance the benefits of early detection against the costs and potential harms of additional imaging.

Standard-of-care risk models, such as the Breast Cancer Risk Assessment Tool^4^ and the Tyrer-Cuzick model,^5^ are based primarily on demographic, reproductive, and clinical risk factors. Although widely used in clinical practice, these models do not directly incorporate imaging-derived features other than breast density, providing limited individualized risk estimation. Breast imaging captures patient-specific structural and textural characteristics of breast tissue that may provide personalized signals for risk prediction.^6,7^ In addition, longitudinal imaging can capture dynamic changes in breast tissue that could signal future cancer development.^8,9^ Recent advances in deep learning have enabled image-based risk prediction models using full-field digital mammography (FFDM).^10–12^ Mirai, as a representative FFDM-based model, has demonstrated improved 5-year risk prediction compared with traditional clinical models.^13^ Additional studies have shown that incorporating longitudinal FFDM exams can further improve predictive performance.^14–16^ However, most existing AI-based risk models rely on two-dimentional projection images,^10–12,14–16^ which may not fully capture three-dimensional structural features relevant to cancer risk.

Digital breast tomosynthesis (DBT) is widely adopted for breast cancer screening in the United States and has largely replaced FFDM in many clinical settings.^17^ DBT generates a quasi three-dimensional representation of the breast from low-dose projections, reducing the masking effect of overlapping fibroglandular tissue.^18^ This allows for a more accurate evaluation of the volumetric density, distribution, and complexity of the breast parenchymal patterns, which have been associated with breast cancer risk.^19–21^ Despite its widespread clinical use in the United States, DBT has been minimally explored for long-term breast cancer risk prediction. Existing research on DBT has primarily focused on cancer detection, short-term risk estimation, or synthetic two-dimensional projections derived from DBT rather than the native three-dimensional volumes.^22–24^ The role of longitudinal DBT for long-term breast cancer risk prediction remains underexplored. Utilizing longitudinal three-dimensional DBT exams may provide complementary structural and temporal signals for risk estimation.

In this retrospective study, we developed and validated a DBT-based risk prediction (DRP) deep learning model that integrates longitudinal DBT exams to estimate individualized 2 to 5-year breast cancer risk. Model performance was evaluated in a large retrospective cohort and compared with an established FFDM-based AI risk model as well as a clinical risk model. The purpose of this study was to assess whether leveraging volumetric and longitudinal imaging information can enable more accurate and personalized long-term breast cancer risk prediction.

## Materials and Methods

This retrospective study was approved by institution A’s Institutional Review Board, and the requirement for informed consent was waived. All patient data were fully de-identified in accordance with the HIPAA guidelines. The study was conducted in compliance with the TRIPOD reporting guidelines.

### Data Collection

The initial data query included consecutive screening and diagnostic DBT exams performed at Institute A between January 1, 2016 and August 31, 2020. Initial search criteria include female patients older than 18 years without breast implants, yielding 392,247 DBT exams from 238,012 women. Secondary exclusions were then applied. Exams without associated radiology reports (*n* = 3,116) were excluded because clinical indications could not be verified. Procedural exams (*n* = 8,510), including biopsy guidance, wire localization, post-procedure imaging, and specimen radiographs, were excluded because those exams were not appropriate for risk prediction.

Following previous work,^14,25^ exams with biopsy-confirmed breast cancer diagnosed within 6 months were excluded to remove prevalent cancers already present at the time of imaging. Finally, exams without any subsequent imaging or pathology follow-up were excluded because cancer risk status could not be reliably determined. The data collection workflow is shown in Figure 1.

**Fig. 1.**
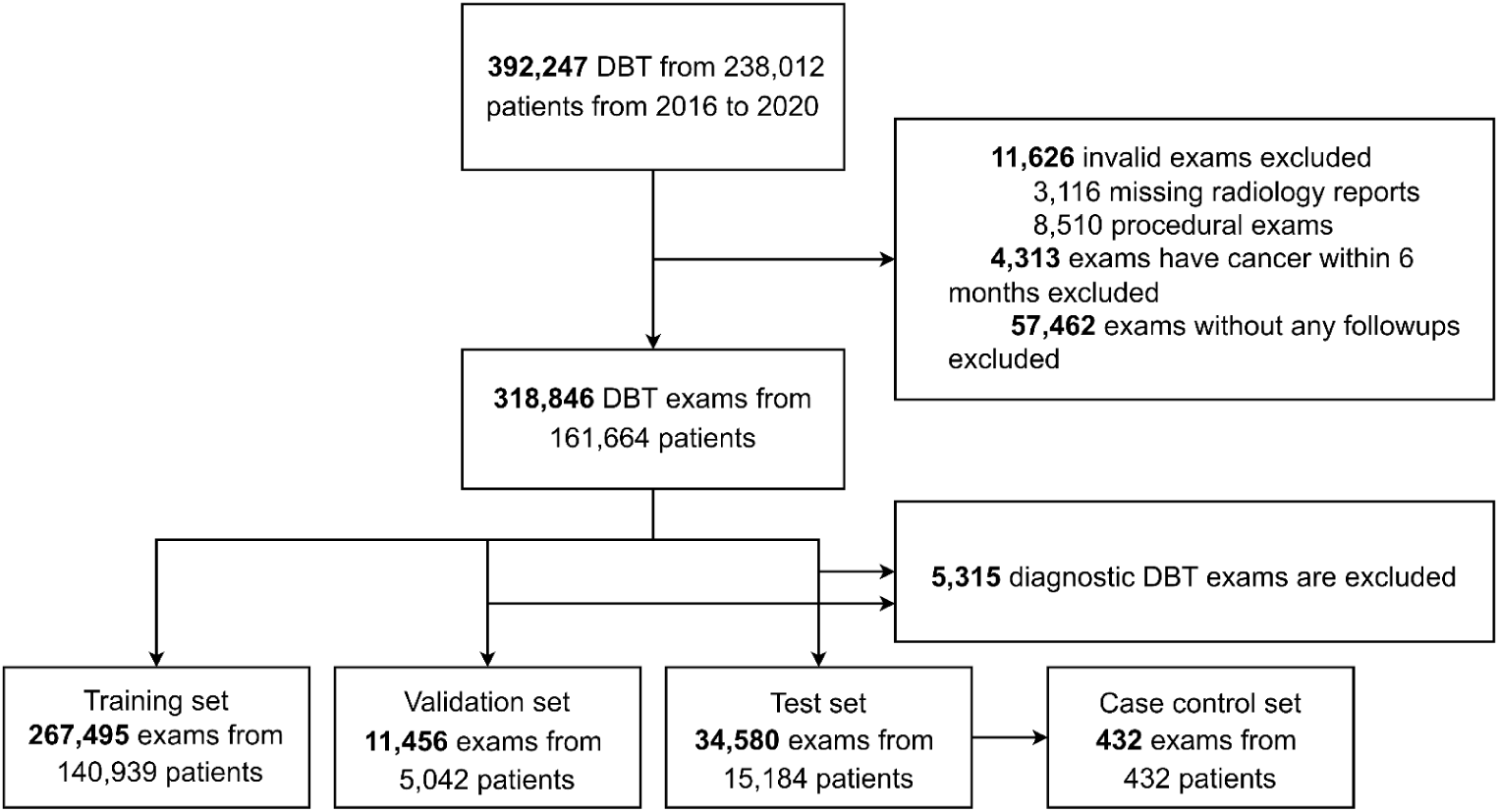
Flow diagram detailing the selection of patients and digital breast tomosynthesis examinations. DBT = digital breast tomosynthesis.

The eligible exams (n = 318,846 exams, 161,664 patients) were randomly split at the patient level into training (n = 267,485 exams, 140,393 patients), validation (n = 11,456 exams, 5,042 patients), and hold-out test (n = 34, 580 exams, 15,184 patients) sets. Diagnostic exams were retained in the training set to maximize data diversity but excluded from the validation and test sets so that final model performance was evaluated on a screening population. In addition, a matched case-control cohort (n = 432) was derived from the test set for comparing against the Tyrer-Cuzick model, consisting of 216 exams from women who were cancer-free at the time of imaging but were diagnosed with breast cancer within 5 years and 216 cancer-negative control exams, with matching performed on age and breast density.

### Cancer Outcome Reference

Breast cancer outcomes were ascertained using pathology records through 2025, and negative outcomes were determined using screening and diagnostic radiology reports through 2025. Malignant outcomes included primary invasive breast cancers, ductal carcinoma in situ, and non-primary breast malignancies, including lymphoma and malignant phyllodes tumors. For each prediction horizon (2–5 years), an exam was labeled cancer-positive if a malignant pathology diagnosis occurred within the corresponding time interval following the exam date. An exam was labeled cancer-negative for a given prediction horizon if no malignant pathology was identified during the entire follow-up period for that horizon and the patient had either only negative imaging results (BI-RADS 1, 2) or positive imaging findings (BI-RADS 0, 3, 4, or 5) that were subsequently confirmed as benign by pathology or the follow-up exam.

### Image Pre-processing

Standard screening and diagnostic mammograms include bilateral FFDM and DBT acquired in the craniocaudal (CC) and mediolateral oblique (MLO) views. Diagnostic DBT exams were obtained using additional targeted views (eg, spot compression and magnification views) as clinically indicated. All DBT exams were acquired using Hologic Selenia Dimensions systems under routine clinical protocols. DBT volumes were exported in their native resolution (4096 × 3328 and 3328 × 2560) with 70 slices on average per volume. Image underwent a standardized pre-processing pipeline.^26^ Non-breast background regions were removed, and a fixed-size window of 2166 × 1339 pixels was cropped to encompass the informative breast regions. All preprocessing steps were applied identically to all exams to preserve longitudinal consistency.

### Model Development

The DRP model is a hierarchical transformer-based deep learning framework designed to predict long-term breast cancer risk from longitudinal DBT exams. Risk was estimated at the time of an index examination using up to 10 prior DBT exams from the same patient. Slice-level features were first extracted using pretrained networks: a global classifier (GMIC)^27^ for global image context and object detectors (YOLOX^28^ and MogaNet^29^) for localized lesion features. These slice-level feature extractors were trained independently to detect breast cancer and were frozen during the subsequent training of the risk model. Slice-level features were then aggregated to generate an exam-level representation using a transformer encoder.^30^ Exam-level representations from all prior and index exams were then combined with clinical variables (age and breast density) and processed by a lightweight two-layer transformer^30^ to model temporal dependencies. The final output was passed through a survival prediction head that estimates the conditional probability of cancer occurrence across discrete yearly intervals (2–5 years after the index exam).^31^ Predicted probabilities were calibrated using Platt scaling,^32^ in which a logistic regression model was fitted on the validation set predictions and applied to the test set. Additional model details are provided in Supplementary Material S1 and Figure S1.

A simplified DRP model using only the index exam was developed to assess the contribution of longitudinal information. In this model, exam-level representations from the index exam, combined with clinical variables, were directly input to the survival prediction head without longitudinal modeling.

### Baseline Risk Model Comparisons

Model performance was compared with two established breast cancer risk assessment methods: a clinical risk model (Tyrer-Cuzick, version 8)^5^ and an FFDM-based deep learning model (Mirai).^11^ Relevant clinical risk factors required for Tyrer-Cuzick calculation were obtained through a structured chart review performed by a trainee (third-year medical student, F.P.) under the supervision of an attending breast radiologist (9 years of experience, L.H.), blinded to outcome status. Tyrer–Cuzick risk scores were calculated using the official Tyrer-Cuzick version 8 software.^33^ Since Mirai generates breast cancer risk estimates from a single 2D FFDM exam, the same-day FFDM exam corresponding to each DBT exam in the test set was used as input to Mirai. Each DBT exam in the dataset was treated as an index exam. All model comparisons were conducted on the same evaluation cohorts using identical performance metrics.

### Risk Stratification Thresholds

We adopted the risk stratification thresholds proposed by prior work^34,35^ based on the NCCN^36^ and ASCO guidelines^37^ to categorize exams into three risk groups based on the predicted 5-year risk. Exams with a predicted risk of less than 1.7% were classified as average risk, those with a predicted risk between 1.7% and 2.999% were classified as increased risk, and those with a predicted risk of 3.0% or higher were classified as high risk.

### Statistical Analysis

Model performance was evaluated using 2-to 5-year areas under receiver operating curves (AUCs), time-dependent concordance index (C-index)^38^, and integrated Brier score (IBS).^39^ The 95% confidence intervals (CIs) were estimated via 10,000 bootstrap samples. For each prediction horizon (2-5 years), exams without sufficient follow-up to confirm cancer-negative status for the corresponding time interval were excluded. Final reported performance reflects an ensemble of the top 5 models with the highest 5-year AUC on the validation set. Comparisons of AUCs were performed using Delong test.^40^ C-index comparisons were conducted using a permutation test^41^ with 10,000 repetitions. Kaplan-Meier curves were compared using the log-rank test.^42^

## Results

The study cohort comprised 161,165 women (mean age, 58.5 ± 11.7 years) who underwent 313,531 DBT exams at Institute A between 2016 and 2020. Among all exams, 2.8% (8,833/313,531) were followed by a breast cancer diagnosis 6 months after the index exam, with an average time to diagnosis of 3.4 ± 1.8 years. The average follow-up among cancer-negative exams was 4.8 ± 1.7 years. Demographic and clinical characteristics are summarized in Table 1.

**Table 1.** Patient and Examination Characteristics for the Institute A Cohort. DBT = digital breast tomosynthesis, SD = standard deviation.

|  | All | Training | Validation | Test | Case-Control |
| --- | --- | --- | --- | --- | --- |
| **Patients, No.** | 161,165 | 140,939 | 5,042 | 15,184 | 432 |
| **Age, mean(SD)** | 58.5 (11.7) | 58.2 (11.8) | 60.8 (11.1) | 60.7 (11.1) | 62.3 (9.08) |
| **Race/Ethnicity, No. (%)** |  |  |  |  |  |
| African American | 16,399 (10.2) | 14,639 (10.4) | 438 (8.7) | 1,322 (8.7) | 26 (6.0) |
| Asian | 7,682 (4.8) | 6,893 (4.9) | 205 (4.1) | 584 (3.8) | 18 (4.2) |
| Hispanic | 2,403 (1.5) | 2,069 (1.5) | 85 (1.7) | 249 (1.6) | 7 (1.6) |
| White | 98,277 (61.0) | 85,053 (60.3) | 3,280 (65.1) | 9,944 (65.5) | 335 (77.5) |
| Other | 6,269 (3.9) | 5,477 (3.9) | 191 (3.8) | 601 (4.0) | 11 (2.5) |
| Unknown | 30,135 (18.7) | 26,808 (19.0) | 843 (16.7) | 2,484 (16.4) | 35 (8.1) |
| **Exams, No.(%)** | 313,531 | 267,495 | 11,456 | 34,580 | 432 |
| **Density** |  |  |  |  |  |
| 1 | 11,316 (7.0) | 9,843 (7.0) | 366 (7.3) | 1,107 (7.3) | 15 (3.4) |
| 2 | 62,389 (38.7) | 54,307 (38.5) | 2,016 (40.0) | 6,066 (39.9) | 165 (36.8) |
| 3 | 75,603 (46.9) | 66,312 (47.1) | 2,317 (46.0) | 6,974 (45.0) | 230 (53.2) |
| 4 | 9,631 (6.0) | 8,525 (6.0) | 280 (5.6) | 826 (5.4) | 28 (6.5) |
| Unknown | 2,226 (1.4) | 1,952 (1.4) | 63 (1.2) | 211 (1.4) | 0 (0.0) |
| **Cancer year, No. (%)** | 8,833 (2.8) | 7,464 (2.8) | 352 (3.1) | 1017 (2.9) | 216 (50%) |
| 6 moths~2 | 2,183 | 1,868 | 78 | 237 | 125 |
| 2~3 | 1,716 | 1,477 | 58 | 181 | 45 |
| 3~4 | 1,603 | 1,353 | 65 | 185 | 36 |
| 4~5 | 1,510 | 1,266 | 66 | 178 | 9 |
| >5 | 1,821 | 1,500 | 85 | 236 | 0 |
| **Time to Cancer, mean** | 3.4 (1.8) | 3.4 (1.8) | 3.6 (1.8) | 3.5 (1.8) | 1.9 (1.0) |
| (SD), year |  |  |  |  |  |
| **Time to latest negative follow-ups, mean (SD), year** | 4.8 (1.7) | 4.7 (1.6) | 5.0 (1.7) | 4.9 (1.7) | 6.1 (0.8) |
| **No. of longitudinal DBTs, mean (SD)** | 5.3 (3.5) | 5.1 (3.5) | 6.0 (3.4) | 6.0 (3.4) | 6.1 (3.4) |
| **Average T-C Score (%), mean (SD)** | - | - | - | - | 7.5 (7.2) |

### Comparison to the Tyrer–Cuzick model

In the matched case-control cohort (n=432), the DRP model achieved a C-index of 0.651 (95% CI, 0.621–0.679) and a 5-year AUC of 0.676 (95% CI, 0.626–0.727), outperforming the Tyrer-Cuzick model, which achieved a C-index of 0.557 (95% CI, 0.520–0.597; p < 0.001) and a 5-year AUC of 0.567 (95% CI, 0.514–0.621; p < 0.001). The DRP model showed higher AUROCs than the Tyrer-Cuzick model across all time horizons (Table 2).

**Table 2.**
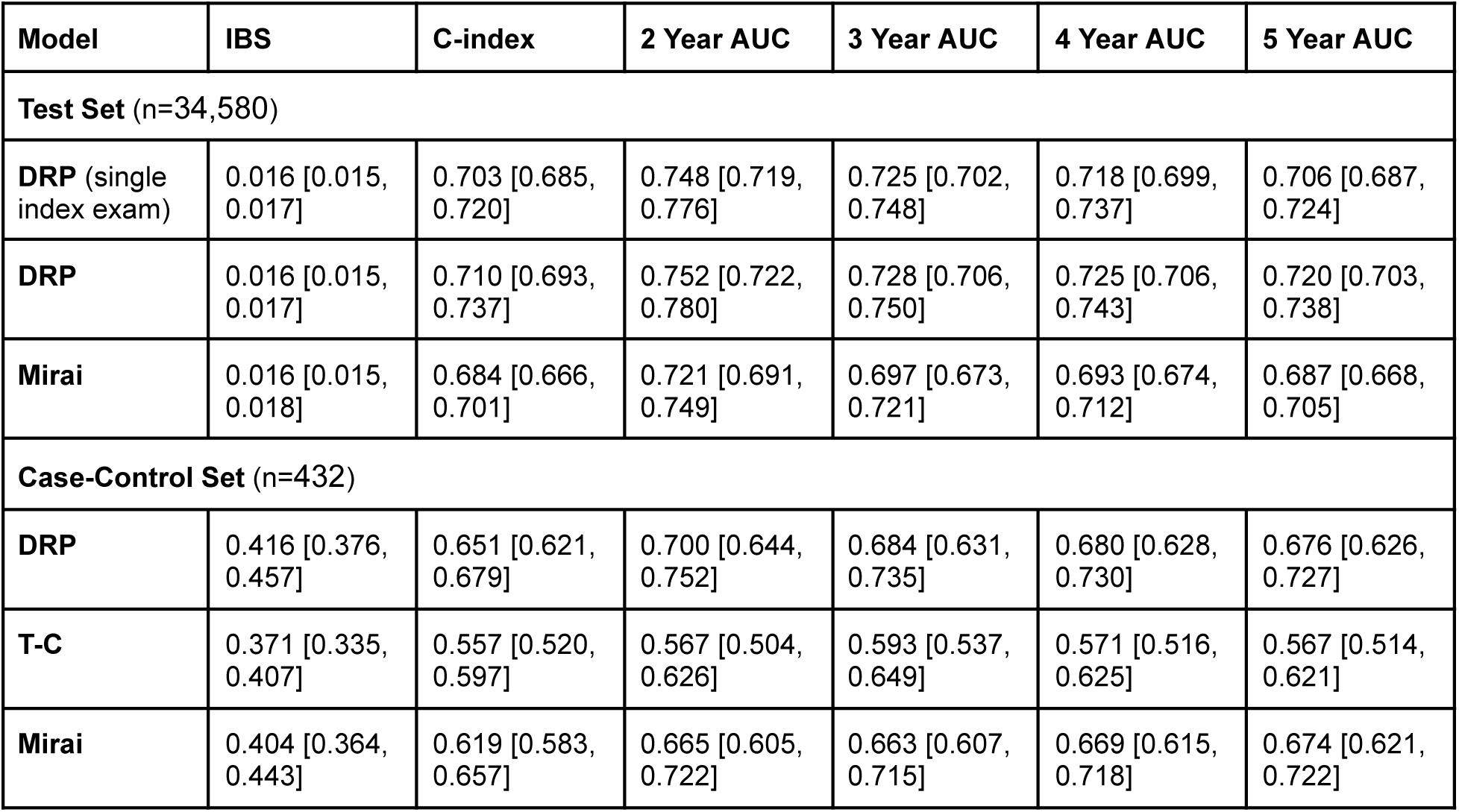
Breast Cancer Risk Prediction Performance. AUC = area under the receiver operating characteristic curve, C-index = concordance index, DRP = deep risk predictor, IBS = integrated Brier score, T-C = Tyrer-Cuzick.

### Long-term Risk Prediction Performance

On the independent hold-out test set (n=34,580), the DRP model using a single index exam achieved a 5-year AUC of 0.706 (95% CI, 0.687–0.724) and a C-index of 0.703 (95% CI, 0.685–0.720). Incorporation of longitudinal DBT exams outperformed DRP that only uses the latest DBT (5-year AUC: 0.720, 95% CI, 0.703–0.738, p < 0.001) and achieved a C-index of 0.710 (95% CI, 0.693–0.737). In comparison, the Mirai model achieved a 5-year AUC of 0.687 (95% CI, 0.668–0.705) and a C-index of 0.684 (95% CI, 0.666–0.701), both of which were lower than those of the longitudinal DRP model (p < 0.001). Improved discrimination of the longitudinal DRP model was observed consistently across 2-to 5-year prediction horizons (Table 2).

Integrated Brier scores were similar between models (DRP, 0.016, 95% CI, 0.015–0.017; Mirai, 0.016, 95% CI, 0.015–0.018). The 5-year calibration curve showed close agreement between predicted and observed cancer incidence across risk strata, with a mean absolute calibration error of 0.005 (Figure 2).

**Figure 2.**
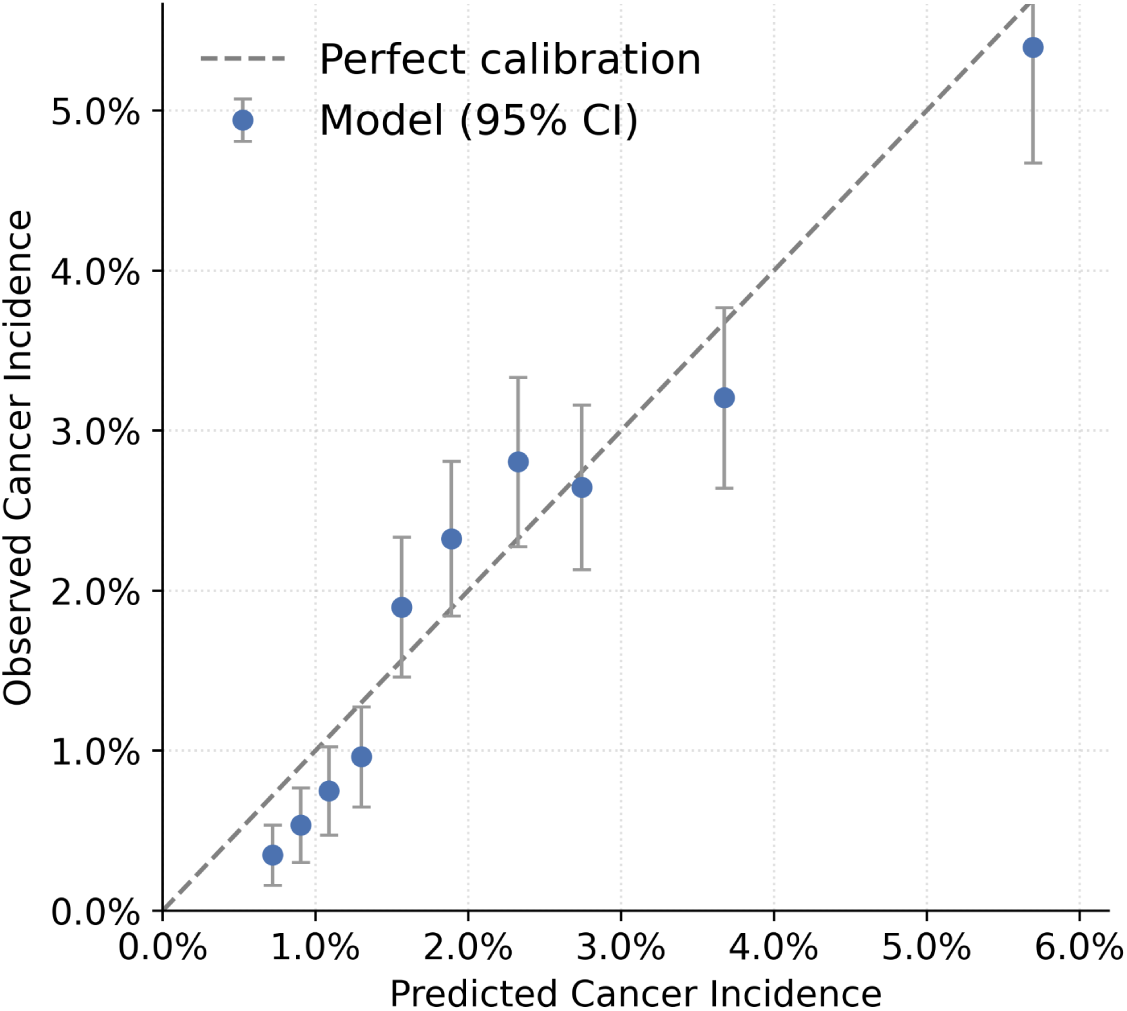
Calibration curve of 5-year breast cancer risk predictions. The graph plots the mean predicted 5-year risk against the observed 5-year cancer incidence across deciles of predicted risk. Error bars represent 95% confidence intervals for the observed incidence, and the dashed diagonal line indicates perfect calibration. The model demonstrates close agreement between predicted and observed risks across the range of estimated probabilities, with a mean absolute calibration error of 0.005.

### Sub-group Analyses

Model performance was evaluated across age, breast density, race/ethnicity, and family history subgroups (Supplementary Table S1). Five-year AUCs across age groups ranged from 0.674 (95% CI, 0.640–0.710) in women aged 60–69 years to 0.749 (95% CI, 0.691–0.803) in women aged 40–49 years. Performance was comparable between women with non-dense and dense breasts, with 5-year AUCs of 0.729 (95% CI, 0.701–0.755) and 0.710 (95% CI, 0.687–0.732), respectively. Across racial and ethnic groups, 5-year AUCs ranged from 0.674 (95% CI, 0.592–0.751) among Asian patients to 0.768 (95% CI, 0.629–0.895) among Hispanic patients. Among patients with and without a family history of breast cancer, 5-year AUCs were 0.689 (95% CI, 0.654–0.722) and 0.728 (95% CI, 0.707–0.749), respectively.

### AI Risk Stratification on High Risk Subgroups

Observed cumulative incidence of cancer and the model’s mean predicted risk across three high-risk phenotype subgroups (age, breast density, and family history) are shown in Figure 3. Kaplan-Meier curves demonstrated significant separation across all three subgroups (log-rank p < 0.001; Figure 3a). The cumulative incidence of cancer increased progressively with older age, higher density, and patients with a positive family history (Figure 3a). Predicted 5-year risk estimates corresponded closely with the observed cumulative incidence across phenotype subgroups (Figure 3b). Predicted risk increased with age, from 1.64% (95% CI, 1.62–1.66%) in women aged 40–49 years to 3.36% (95% CI, 3.32–3.40%) in women aged ≥70 years (p < 0.001). Women with dense breasts (BI-RADS C–D) had a higher mean predicted risk (2.57%; 95% CI, 2.54–2.59%) than those with non-dense breasts (BI-RADS A–B) (2.31%; 95% CI, 2.28–2.33%) (p < 0.001). Women with a first-degree family history had a higher predicted risk (2.62%; 95% CI, 2.58–2.65%) than those without (2.39%; 95% CI, 2.37–2.41%) (p < 0.001).

**Fig. 3.**
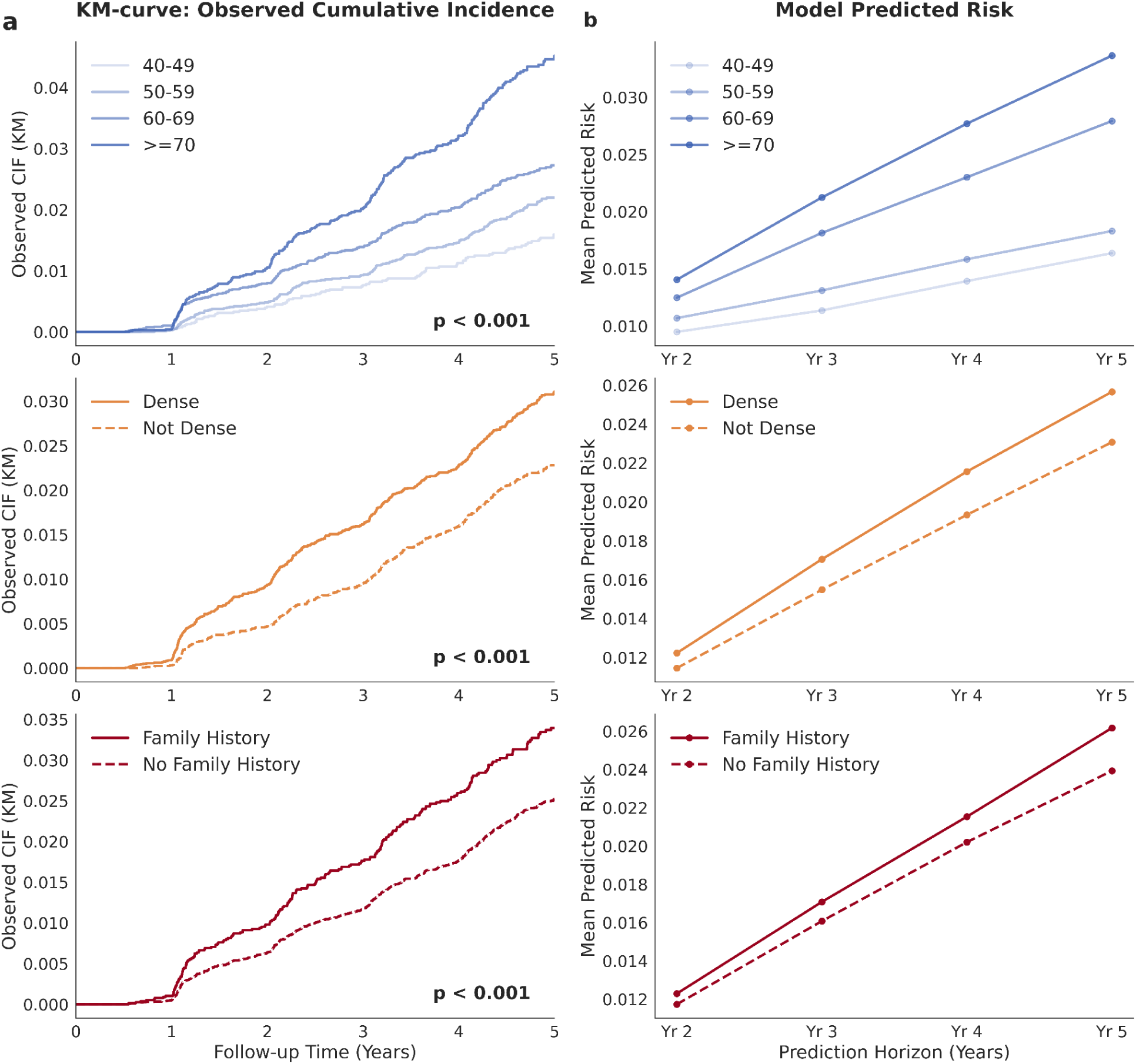
Observed cumulative incidence versus model-predicted breast cancer risk. (A) Kaplan-Meier cumulative incidence curves stratified by age group, breast density (BI-RADS categories C and D vs A and B), and the presence or absence of a first-degree family history of breast cancer. (B) Mean predicted breast cancer risk from 2 to 5 years across the corresponding phenotype groups. The model’s predicted risk trajectories showed alignment with observed cumulative incidence across all evaluated demographic and clinical subgroups. BI-RADS = Breast Imaging Reporting and Data System.

### Risk Stratification among Breast Density Groups

Table 3 summarizes the observed 5-year cancer incidence of DRP-based risk stratification across breast density categories. The proportion of women classified as high risk was higher among women with dense breasts (BI-RADS C–D) (C: 28.4%, 4,476/15,770; D: 26.6%, 500/1,877) than among those with non-dense breasts (BI-RADS A–B) (A: 15.5%, 404/2065; B: 25.0%, 3,459/13,853), consistent with the higher overall cancer incidence observed in denser breasts. Within each density category, the DRP model further stratified women into distinct risk groups. Across all density groups, women classified as average risk consistently demonstrated the lowest 5-year cancer incidence (0.6%–0.9%). In fatty, scattered, and heterogeneously dense breasts (BI-RADS A–C), women classified as high risk had higher cancer incidence than those classified as increased-risk. In extremely dense breasts (BI-RADS D), cancer incidence in the high-risk group (3.0%, 15/500) was slightly lower than that observed in the increased-risk group (3.7%, 25/672).

**Table 3.** AI-Assigned 5-Year Breast Cancer Risk and Incidence by BI-RADS Breast Density Category. Risk groups are defined by 5-year risk thresholds: high (≥3.0%), increased (1.7%–2.9%), and average (<1.7%). BI-RADS = Breast Imaging Reporting and Data System.

| Breast Density<br>N (%) | 5-Year Cancer Incidence | AI Risk Category | % within Density Group | 5-Year Cancer Incidence |
| --- | --- | --- | --- | --- |
| A – Fatty<br>2,605 (7.5 %) | 1.3 % | High ($\geq 3.0\%$) | 15.5% (404/ 2,605) | 2.5% (10/404) |
|  |  | Increased (1.7-2.9%) | 22.8% (595/ 2,605) | 2.2% (13/595) |
| | | Average ($<1.7\%$) | 61.7% (1,606/2,605) | 0.6% (10/1,606) |
| B – Scattered<br>13,853 (40.1 %) | 2.0 % | High ($\geq 3.0\%$) | 25.0% (3,459/13,853) | 4.0% (137/3,459) |
|  |  | Increased (1.7-2.9%) | 28.7% (3,977/13,853) | 2.2% (88/3,977) |
| | | Average ($<1.7\%$) | 46.3% (6,417/13,853) | 0.7% (48/6,417) |
| C – Hetero Dense<br>15,770 (45.6 %) | 2.6 % | High ($\geq 3.0\%$) | 28.4% (4,476/15,770) | 4.9% (220/4,476) |
|  |  | Increased (1.7-2.9%) | 32.5% (5,120/15,770) | 2.6% (133/5,120) |
| | | Average ($<1.7\%$) | 39.2% (6,174/15,770) | 0.9% (56/6,174) |
| D – Extremely Dense<br>1,877 (5.4 %) | 2.4 % | High ($\geq 3.0\%$) | 26.6% (500/1,877) | 3.0% (15/500) |
|  |  | Increased (1.7-2.9%) | 35.8% (672/1,877) | 3.7% (25/672) |
| | | Average ($<1.7\%$) | 37.6% (705/1,877) | 0.7% (5/705) |

### Comparison of Risk Stratification by DRP and Mirai

We examined breast cancer incidence rate among women stratified into each risk category based on the DRP and Mirai (Figure 4, Table 4). In the high-risk category (≥3.0% predicted 5-year risk), women classified as high risk by both models had the highest observed incidence (5.2%; 253/4,895; 95% CI: 4.57–5.83). Among cases where only one model assigned high risk, incidence was 3.4% (147/4,268; 95% CI, 2.92–4.04) for those categorized as high risk by the DRP model only, compared with 3.0% (93/3,087; 95% CI, 2.44–3.68) for those categorized as high risk by Mirai only. In the average-risk group (<1.7% predicted 5-year risk), cases classified as average risk by both models had the lowest observed incidence (0.5%; 44/8,735; 95% CI, 0.37–0.68). Among cases where only one model assigned average risk, incidence was 1.2% (76/6,204; 95% CI, 0.97–1.53) for DRP only and 1.5% (46/3,019; 95% CI, 1.12–2.03) for Mirai only.

**Fig. 4.**
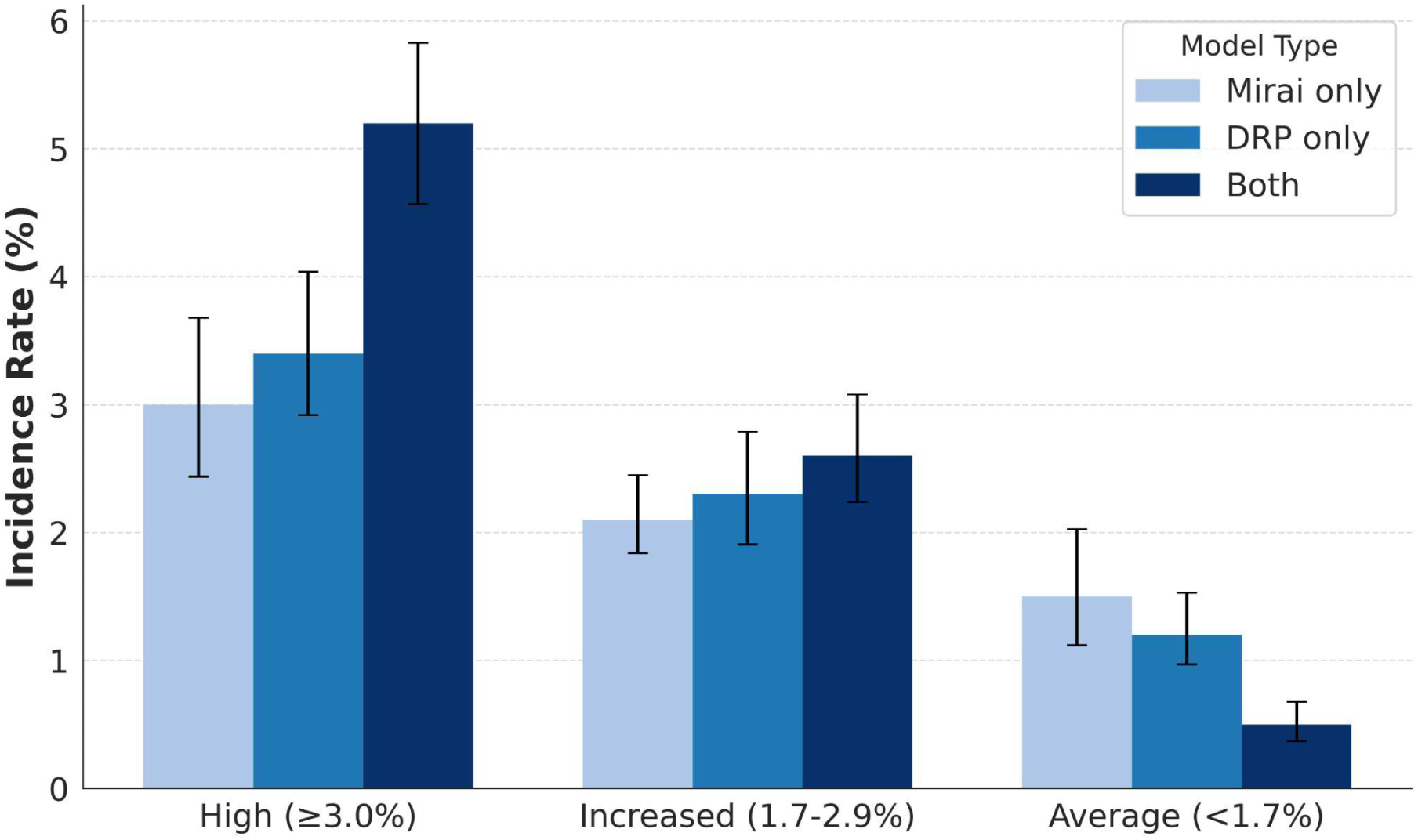
Comparison of 5-year breast cancer incidence across risk stratifications assigned by the DRP and Mirai models. Bar graphs display the observed incidence for examinations classified into high (≥ 3.0%), increased (1.7%–2.9%), and average (< 1.7%) predicted 5-year risk categories. The bars distinguish between concordant assignments (exams assigned to the same category by both models) and discordant assignments (assigned by only one model). DRP = DBT-based risk prediction model.

**Table 4.** Comparison of 5-Year Breast Cancer Incidence Between Concordant and Discordant Risk Classifications by the DBT-based risk prediction model (DRP) and Mirai. Risk groups are defined as high (≥3.0%), increased (1.7%–2.9%), and average (<1.7%) 5-year risk. “Both” indicates exams assigned to the same risk category by both models; “Mirai only” and “DRP only” indicate discordant classifications.

| AI Risk Category | Model | N (Total) | Cancer Cases | Incidence Rate (%) | 95% CI |
| --- | --- | --- | --- | --- | --- |
| High ($\geq 3.0\%$) | Mirai only | 3,087 | 93 | 3.0% | (2.44 - 3.68) |
|  | DRP only | 4,268 | 147 | 3.4% | (2.92 - 4.04) |
|  | Both | 4,895 | 253 | 5.2% | (4.57 - 5.83) |
| Increased ($1.7 - 2.9\%$) | Mirai only | 9,064 | 193 | 2.1% | (1.84 - 2.45) |
|  | DRP only | 4,698 | 109 | 2.3% | (1.91 - 2.79) |
|  | Both | 5,772 | 152 | 2.6% | (2.24 - 3.08) |
| Average ($< 1.7\%$) | Mirai only | 3,019 | 46 | 1.5% | (1.12 - 2.03) |
|  | DRP only | 6,204 | 76 | 1.2% | (0.97 - 1.53) |
|  | Both | 8,735 | 44 | 0.5% | (0.37 - 0.68) |

## Discussion

In this retrospective study of more than 160,000 women undergoing DBT, we developed and evaluated the DRP model, a deep learning model that integrates longitudinal DBT exams, to estimate 2–5-year breast cancer risk. The model demonstrated improved discrimination performance compared with the FFDM-based Mirai model in the independent hold-out test set (AUC, 0.720 vs 0.687; p < 0.001) and the Tyrer-Cuzick model in the matched case-control cohort (AUC, 0.676 vs 0.567; p < 0.001). The predicted risk was aligned with increasing observed cancer incidence across subgroups defined by age, breast density, and family history, indicating consistent risk stratification with established risk factors. Moreover, the model provides refined risk stratification within breast density categories, identifying heterogeneity in risk beyond density alone. These findings suggest the utility of longitudinal DBT for personalized, long-term breast cancer risk assessment beyond categorical risk factors.

The DRP model builds upon prior work on DBT-based breast cancer risk prediction. Eriksson et al. used DBT-derived mammographic features instead of full images for interval cancer prediction (1–2 year risk prediction), reporting AUCs of 0.82^22^ and 0.72^22,43^ in a subsequent validation. Jiang et al. developed a model on synthetic 2D reconstructions derived from DBT for long-term prediction, achieving an externally validated 5-year AUC of 0.72.^23^ More recently, Dorster et al. trained a model directly on 3D DBT volumes and reported a 5-year AUROC of 0.80^44^ but their approach did not incorporate longitudinal exams. In contrast, our model directly analyzes longitudinal native 3D DBT volumes. Incorporating multiple exams improved performance over a single-time point model (AUC, 0.720 vs 0.706; p < 0.001), suggesting that temporal information provides additional predictive value.

The improved performance of DBT-based models over the FFDM-based model may reflect enhanced volumetric detail captured by tomosynthesis. Prior studies have shown that mammographic texture captures features associated with breast cancer risk, including associations with genetic markers,^6,7,45^ much of which is not fully accounted for by traditional clinical risk factors.^46^ By providing volumetric data, DBT enables more comprehensive characterization of parenchymal structure than FFDM, making its derived radiomic and texture features more strongly associated with risk estimation.^47,48^ In addition, evaluating multiple exams over time captures evolving imaging patterns. Changes in breast density and texture reflect shifts in hormones, environment, and lifestyle^49,50^ and may provide predictive information beyond that available from single-timepoint models.^8,9^ At last, our joint risk stratification using both DRP and Mirai further suggested that FFDM and DBT may provide complementary information for breast cancer risk assessment.

The model stratified risk independent of breast density, identifying substantial heterogeneity in cancer risk within each density category. It reclassified over one-third of women with extremely dense breasts (37.6%, 705/1,877) as average risk, corresponding to a 0.7% (5/705) cancer incidence. Conversely, the model identified a high-risk subset among women with fatty breasts (15.5%, 404/2,605) who demonstrated a cancer incidence of 2.5% (10/404). These findings highlight the limitations of breast density as a standalone risk indicator and demonstrate the potential of AI-based imaging models to refine screening decisions beyond density. In clinical practice, this approach may help inform decisions regarding supplemental imaging and screening intensity, i.e. reducing unnecessary supplemental imaging among women with dense breasts while identifying high-risk individuals who may benefit from intensified screening or preventive strategies. Furthermore, integration of automated risk assessment at the point of care may further facilitate implementation of risk-adapted screening, addressing challenges observed in prior trials where uptake was limited by the need for additional genetic testing or clinical data collection,^3^ although prospective validation is needed to determine its impact on clinical outcomes and adherence. This approach aligns with emerging screening paradigms that incorporate individualized risk estimates to guide screening strategies, with increasing interest in the role of imaging-derived features.^3,51,52^

This study has several limitations. First, the model was developed and retrospectively validated using data from a single academic health system, which may limit generalizability to other populations and practice settings. Although the cohort included both academic and community sites, external validation and prospective evaluation are needed to further assess real-world performance. Second, most DBT exams in our dataset were acquired on Hologic systems, which represent the majority of the U.S. market but may not reflect performance across other vendors or imaging protocols. Third, this version of the model did not incorporate established risk factors including genetic information, reproductive history, additional clinical history and findings from other imaging modalities (e.g., MRI or ultrasound), which may further improve individualized risk estimation. Finally, model performance was lower in Asian women compared to other subgroups, potentially reflecting underrepresentation in the training and validation cohort; validation in larger and more diverse populations is warranted.

In conclusion, this study demonstrates that the use of longitudinal DBT exams enables more accurate personalized long-term breast cancer risk prediction. These findings suggest that longitudinal DBT provides complementary volumetric and temporal information beyond existing clinical and FFDM-based approaches. Future work should focus on external validation across diverse institutions and vendors, integration of a wider range of risk factors, as well as prospective studies to evaluate the role of DBT-based risk prediction in personalized breast cancer screening strategies.

## Data Availability

Data analyzed during the study were provided by a third party. Requests for data should be directed to the provider indicated in the Acknowledgements.

