## Supplementary Materials for "Predicting 5-Year Breast Cancer Risk from Longitudinal Digital Breast Tomosynthesis: A Single-center Retrospective Study"

**Model Architecture**
We developed a four-stage hierarchical transformer architecture to progressively aggregate slice, exam, and longitudinal-level information from index and prior DBT exams into a unified representation for long-term risk prediction.

**Stage 1: Slice-Level Feature Extraction.**
To capture complementary image representations, we used two types of pre-trained, frozen feature extractors: a classifier network (GMIC)[27] for global embeddings and object detectors (YOLOX[28] and MogaNet[29]) for localized lesion features. Feature extraction was performed at the 2D slice level across all prior and index DBT exams. During model training, two slices per DBT view were uniformly sampled from the top 10 most informative slices, as ranked by the extractors' prediction scores. During inference, features were extracted from all slices in each DBT volume. This ensures that the complete 3D volumetric and structural information of the breast is fully preserved and utilized for the final risk prediction.

**Stage 2. Exam-level Representation Learning**
For each DBT exam, slice-level embeddings from all views were aggregated to form an exam-level representation. Detector-derived features were concatenated with 32-dimensional learnable embeddings encoding imaging metadata, including laterality (left/right), view type (e.g., CC, MLO), and bounding box positions. The combined sequence of slice-level tokens were processed by a DeiT-based transformer encoder,[53] which models relationships across slices and views. The output cls token is the exam-level representation of the DBT exam.

**Stage 3. Longitudinal Representation Learning**
Exam-level embeddings from all available time points (including the index and prior examinations) were ordered chronologically and concatenated with clinical variables, including patient age, breast density, and examination date. A two-layer transformer encoder processed this sequence to model temporal dependencies and parenchymal changes over time.

**Stage 4. Survival Head for Risk Prediction**
The lightweight transformer output was passed through a survival prediction head that estimated the conditional hazard probability of developing breast cancer within each yearly discrete time interval (2–5 years). The prediction head is a multi-layer perceptron. Risk was modeled using a discrete-time survival framework, in which the network predicts the probability of event occurrence within each interval conditional on prior survival.[31] The model was trained using a discrete-time survival loss based on the negative log-likelihood of the observed event times.

### Supplementary Figures and Tables

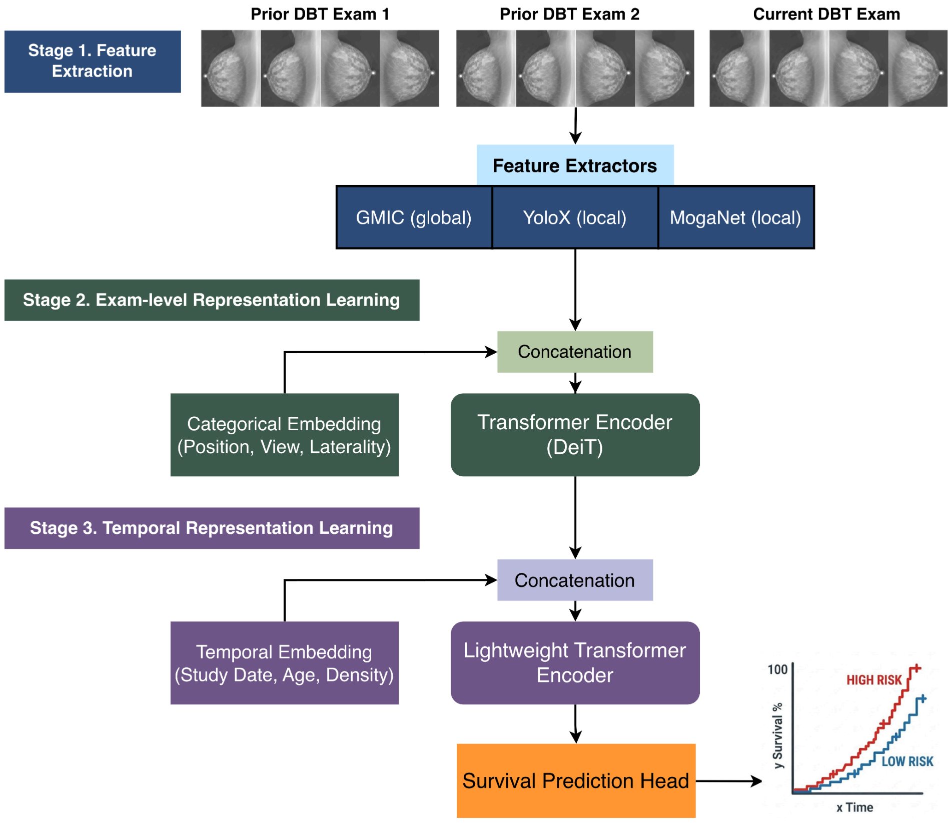


**Figure S1. Overview of the DBT-based Risk Prediction (DRP) model architecture.** In stage 1, prior and index DBT examinations are processed through pre-trained feature extractors (GMIC, YOLOX, and MogaNet) to capture complementary imaging representations. In stage 2, extracted features from multiple views are concatenated and encoded via a transformer module to generate a unified examination-level representation. In stage 3, representations from sequential DBT examinations are combined with temporal and clinical embeddings (examination date, age, and breast density) and processed by a lightweight transformer encoder to model longitudinal parenchymal changes. In stage 4, the resulting temporal representation is passed to a survival prediction head to estimate individualized breast cancer risk over time. DBT = digital breast tomosynthesis, DRP = DBT-based risk prediction.

**Supplementary Table S1. The 5 year risk prediction performance within each subgroup.** Model performance is reported within each demographic and density subgroup. For breast density, BI-RADS A-B are classified as Not Dense, and BIRADS C-D are classified as Dense. Family history includes first degree family member's history of breast cancer.

| Subgroups | IBS | C-index | 2 Year AUC | 3 Year AUC | 4 Year AUC | 5 Year AUC |
|---|---|---|---|---|---|---|
| **Age** | | | | | | |
| 40~49 | 0.009 [0.007, 0.012] | 0.737 [0.682, 0.790] | 0.815 [0.729, 0.892] | 0.743 [0.668, 0.814] | 0.734 [0.668, 0.796] | 0.749 [0.691, 0.803] |
| 50~59 | 0.012 [0.010, 0.014] | 0.716 [0.683, 0.747] | 0.788 [0.729, 0.843] | 0.729 [0.682, 0.773] | 0.729 [0.693, 0.764] | 0.717 [0.683, 0.750] |
| 60~69 | 0.017 [0.015, 0.019] | 0.670 [0.638, 0.703] | 0.698 [0.643, 0.752] | 0.692 [0.649, 0.734] | 0.690 [0.655, 0.727] | 0.674 [0.640, 0.710] |
| >= 70 | 0.026 [0.022, 0.029] | 0.669 [0.636, 0.701] | 0.686 [0.627, 0.744] | 0.686 [0.641, 0.729] | 0.676 [0.639, 0.713] | 0.679 [0.644, 0.713] |
| **Density** | | | | | | |
| Not Dense | 0.013 [0.011, 0.014] | 0.722 [0.697, 0.747] | 0.754 [0.704, 0.802] | 0.740 [0.702, 0.776] | 0.736 [0.707, 0.764] | 0.729 [0.701, 0.755] |
| Dense | 0.019 [0.017, 0.021] | 0.695 [0.673, 0.717] | 0.734 [0.696, 0.769] | 0.712 [0.682, 0.741] | 0.711 [0.687, 0.736] | 0.710 [0.687, 0.732] |
| **Race/Ethnicity** | | | | | | |
| African American | 0.014 [0.010, 0.018] | 0.707 [0.653, 0.758] | 0.710 [0.602, 0.809] | 0.728 [0.641, 0.808] | 0.722 [0.650, 0.788] | 0.706 [0.645, 0.764] |
| Asian | 0.027 [0.019, 0.036] | 0.665 [0.595, 0.732] | 0.645 [0.496, 0.783] | 0.659 [0.565, 0.743] | 0.680 [0.603, 0.752] | 0.674 [0.592, 0.751] |
| Hispanic | 0.012 [0.005, 0.022] | 0.739 [0.580, 0.872] | 0.889 [0.745, 0.972] | 0.875 [0.755, 0.968] | 0.799 [0.648, 0.926] | 0.768 [0.629, 0.895] |
| White | 0.018 [0.016, 0.019] | 0.711 [0.691, 0.731] | 0.752 [0.719, 0.783] | 0.729 [0.703, 0.756] | 0.726 [0.705, 0.748] | 0.723 [0.703, 0.743] |
| **Family History** | | | | | | |
| Yes | 0.021 [0.018, 0.024] | 0.678 [0.647, 0.708] | 0.710 [0.660, 0.757] | 0.677 [0.637, 0.716] | 0.685 [0.651, 0.718] | 0.689 [0.654, 0.722] |
| No | 0.015 [0.013, 0.016] | 0.720 [0.700, 0.739] | 0.761 [0.725, 0.795] | 0.746 [0.718, 0.773] | 0.738 [0.715, 0.760] | 0.728 [0.707, 0.749] |
